# Longitudinal Fecal Shedding of SARS-CoV-2, Pepper Mild Mottle Virus, and Human Mitochondrial DNA in COVID-19 Patients

**DOI:** 10.1101/2024.04.22.24305845

**Authors:** Pengbo Liu, Orlando III Sablon, Yuke Wang, Stephen Patrick Hilton, Lana Khalil, Jessica Mae Ingersoll, Jennifer Truell, Sri Edupuganti, Ghina Alaaeddine, Amal Naji, Eduardo Monarrez, Marlene Wolfe, Nadine Rouphael, Colleen Kraft, Christine L. Moe

**Affiliations:** Center for Global Safe Water, Sanitation, and Hygiene, Rollins School of Public Health, Emory University, Atlanta, Georgia, USA; Hope Clinic of the Emory Vaccine Center, Division of Infectious Diseases, Department of Medicine, School of Medicine, Emory University, Atlanta, Georgia, USA; Division of Infectious Diseases, Emory University School of Medicine, Atlanta, Georgia, USA

**Keywords:** SARS-CoV-2, COVID-19, longitudinal, fecal shedding, PMMoV, mtDNA

## Abstract

Since the coronavirus disease 2019 (COVID-19) pandemic, wastewater-based epidemiology (WBE) has been widely applied in many countries and regions for monitoring COVID-19 transmission in the population through testing severe acute respiratory syndrome coronavirus 2 (SARS-CoV-2) in wastewater. However, the lack of dynamic level of viral shedding in the wastewater and accurate number of infections in the community creates challenges in predicting COVID-19 prevalence in the population and interpreting WBE results. In this study, we measured SARS-CoV-2, pepper mild mottle virus (PMMoV), and human mitochondrial DNA (mtDNA) in longitudinal fecal samples collected from 42 COVID-19 patients for up to 42 days after diagnosis. SARS-CoV-2 RNA was detected in 73.1% (19/26) of inpatient study participants in at least one of the collected fecal specimens during the sampling period. Most participants shed the virus within three weeks after diagnosis, but five inpatient participants still shed the virus between 20 and 60 days after diagnosis. The median concentration of SARS-CoV-2 in positive fecal samples was 1.08×10^5^ genome copies (GC)/gram dry fecal material. PMMoV and mtDNA were detected in 99.4% (154/155) and 100% (155/155) of all fecal samples, respectively. The median concentrations of PMMoV RNA and mtDNA in fecal samples were 1.73×10^7^ and 2.49×10^8^ GC/dry gram, respectively. These results provide important information about the dynamics of fecal shedding of SARS-CoV-2 and two human fecal indicators in COVID-19 patients. mtDNA showed higher positive rates, higher concentrations, and less variability between and within individuals than PMMoV, suggesting that mtDNA could be a better normalization factor for WBE results than PMMoV.

## 1. INTRODUCTION

COVID-19 (coronavirus disease 2019) is caused by SARS-CoV-2 (severe acute respiratory coronavirus 2), a positive-sense single-stranded RNA virus. Although COVID-19 is a respiratory disease and patients predominantly manifest respiratory symptoms, systemic and respiratory manifestations are often accompanied by gastrointestinal symptoms and fecal shedding of viral RNA. Meta-analysis of studies that focus on gastrointestinal (GI) infection and fecal shedding in patients with COVID-19 report the prevalence of GI symptoms to be around 17.6% (1) and 28.5% (2) in patients with severe COVID-19 during the acute infection phase. In COVID-19 patients, the prevalence of positive RNA in stool samples was about 50% (1, 3) within the first week of diagnosis. Interestingly, several studies that have collected paired longitudinal respiratory and fecal samples demonstrated prolonged fecal shedding and higher viral load in feces compared to the paired respiratory samples collected at the same time period (3, 4). This suggests that SARS-CoV-2 infection of the GI tract persists longer than in the respiratory tract and the GI tract excretes more viruses than the respiratory tract. SARS-CoV-2 concentration in stool was reported to be between 10^2^ to 10^7^ GC/mL in an early study (5) for the prototype SARS-CoV-2, but there is a lack of this information for the recent variants.

While the presence and prevalence of SARS-CoV-2 RNA in the GI tract are well established, much less is known about the duration and the amount of SARS-CoV-2 fecal shedding in patients with COVID-19. Natarajan et al. (3) reported that 12.7% of subjects still shed SARS-CoV-2 at 4 months and 3.8% of patients shed SARS-CoV-2 at 7 months post diagnosis. Whereas there was no evidence of ongoing oropharyngeal virus shedding at this time. Additionally, one study suggested that fecal SARS-CoV-2 RNA shedding may start 3-4 days before the onset of the symptoms (5), indicating that the duration of SARS-CoV-2 fecal shedding may be even longer.

Many studies have observed significant correlations between the SARS-CoV-2 RNA concentration in wastewater and the number of confirmed COVID-19 cases reported in the corresponding wastewater catchment areas (6–12), which highlights the application of WBE to monitor the burden of viral infection in the population. Other studies have attempted to use SARS-CoV-2 fecal shedding levels as parameters in models to predict the prevalence or incidence of COVID-19 in communities (8, 13, 14). Predicting prevalence or incidence in the population is critical for public health decision-making.

This application has been explored via several statistical modeling approaches (14–16); However, using modeling to accurately predict COVID-19 prevalence or incidence in populations from SARS-CoV-2 concentrations in wastewater requires longitudinal and quantitative SARS-CoV-2 fecal shedding data. To date, there have been several reports on SARS-CoV-2 fecal shedding, but many studies lack longitudinal samples, quantitative SARS-CoV-2 concentrations (report Ct values rather than absolute quantification) in stool, accurate timing of when the samples were collected after the onset of symptoms, specific amount of fecal material used for viral quantification, and the limit of detection of the assay used for viral quantification, etc. This missing information in fecal shedding data limits the appropriate application of the models and the interpretation of WBE modeling results.

Pepper mild mottle virus (PMMoV) and human mitochondrial DNA (mtDNA) are two human fecal indicators which are often analyzed along with SARS-CoV-2 quantification in wastewater samples, and both are used for normalization of SARS-CoV-2 concentrations in wastewater. PMMoV, a plant RNA virus that causes infection in pepper crops, is excreted in high concentrations in human fecal material if people consume food products containing infected peppers. Up to 10^9^ virion particles of PMMoV have been reported per gram of human feces by dry weight (17). Because of its abundance in human stool, persistence in the environment, and multiple exposure pathways to humans, PMMoV is frequently reported as a human fecal indicator in WBE studies (18, 19). Human mtDNA is exclusively human in origin and highly abundant in human feces. However, it is less frequently reported (20, 21) than PMMoV and is potentially a new, reliable human fecal indicator. This marker has been detected in high copy numbers in 100% of human fecal specimens across multiple geographic regions (22) and in 92% of sewage samples (23).

The objective of this study was to examine the fecal shedding dynamics of SARS-CoV-2 RNA, PMMoV, and mtDNA in longitudinal fecal samples collected from confirmed COVID-19 inpatient and outpatient participants. The results can be used to better predict COVID-19 prevalence or incidence in communities using mathematical modeling and guide interpretation of SARS-CoV-2 RNA levels in wastewater samples using WBE approaches.

## 2. MATERIALS and METHODS

### 2.1. Study Description and Stool Sample Collection

Study participants were recruited from the inpatient clinic at Emory University Hospital between March 21, 2021 and September 21, 2021 and the outpatient clinic by the Hope Clinic of Emory Vaccine Center between September 22, 2021 and July 28 2022. All participants were required to sign an informed consent. In addition, information on demographics, clinical symptoms, and COVID-19 vaccination was also collected.

COVID-19 patients were enrolled as study participants if an in-house real-time RT-PCR test detected SARS-CoV-2 RNA in their nasopharyngeal swab samples or other commercial assays (e.g., antigen test) showed SARS-CoV-2 antigen in their saliva specimens within 7 days after the onset of COVID-19 symptoms. Participants with any behavioral, cognitive, or psychiatric condition were excluded from the enrollment. The day with SARS-CoV-2 positive detection was considered the COVID-19 confirmation date in this study. The first stool sample was collected after the participant was enrolled, usually within 7 days after the confirmation date. The date of the first stool sample collection was defined as Day 1, and subsequent samples were collected on Days 3, 7, 14, 28, and 42 from the first stool sample for each subject. The sampling day was converted to the day of COVID-19 confirmation for data analysis and visualization purposes. Stool collection kits were provided to study participants. For inpatients, stool samples were picked up from the hospital room and there was usually no delay. For outpatients, stool collection kits were shipped to the home address, and study participants were asked to ship samples back to the Emory study lab using prepaid mailers. These samples were usually delayed for shipment and may not have been kept all the way in the cold chain. Once received by the study lab, the samples were immediately stored at -80℃ prior to nucleic acid extraction and RT-qPCR detection.

### 2.2. Fecal Sample Processing and Nucleic Acid Extraction

Stool samples were processed by the physical disruption method of bead lysing followed by total nucleic acid extraction with an automated purification platform, Qiagen EZ1 Advanced XL (Qiagen, #9001875). First, fecal specimens were thawed on ice from -80°C. Each specimen was precisely weighed on an analytical balance, ranging between 27.0 mg to 33.1 mg, into a 2.0 mL pre-filled 0.7 mm garnet bead tube (Omni International, #19-624). Then, 600 µL of Qiagen PowerBead solution (Qiagen, #12955-4-BS) was added to each wet sample. Specimens were immediately mixed on a Bead Genie (Scientific Industries Inc, #SI-B100) at 3,000 rpm for 2 minutes, which introduces physical agitation resulting in the lysing of the sample matrix and homogenizing the sample. A 200 µL aliquot of lysate was added to 5,000 genome copies (10 µL) of Bovine Respiratory Syncytial Virus (BRSV) (INFORCE 3, Zoetis, Parsippany, NJ) and processed by the Qiagen EZ1 Advanced XL with the Qiagen EZ1 DSP virus kit (Qiagen, # 62724) and Qiagen EZ1 Advanced DSP Virus Card (Qiagen, #9018306). BRSV served as an internal processing control during the concentration and extraction procedures. Total nucleic acids were extracted by a Zymo Research OneStep™ PCR Inhibitor Removal kit (Zymo Research, #D6030). A Zymo-Spin^TM^ III-HRC Column (Zyme Research, #C1005) was used for each sample as per the manufacturer’s instructions. We inserted the column into a collection tube and added 600 μL of Prep-Solution. Then, we centrifuged the column at 8,000 × g for 3 minutes. The prepared column was transferred to a clean 1.5 ml microcentrifuge tube, and 60 μL of RNA was added to the Zymo-Spin^TM^ III-HRC Column. The sample was then centrifuged at 16,000 × g for 3 minutes. Total nucleic acid was collected and stored at 80°C.To estimate the dry weight, each fecal sample was weighed, placed onto an aluminum weigh boat, incubated at 105-110°C for 24 hours, and re-weighed after incubation.

The dry weight percent for the calculation of SARS-CoV-2 concentrations per mass of dry weight was calculated using the following formula:

[1-(wet mass of fecal material - dry mass of fecal material)/wet mass of fecal material] ×100%

### 2.3. Quantification of SARS-CoV-2, PMMoV, and mtDNA in Stool Samples using dPCR

Digital PCR was performed using the QIAcuity Digital PCR System and QIAcuity OneStep Advanced Probe Kit (Qiagen, #250132) following the manufacturer’s protocol. The Qiagen QIAcuity instrument was programmed using the following parameters.

Reverse transcription was set at 50°C for 40 minutes for 1 cycle followed by PCR initial heat activation at 95°C for 2 minutes. The PCR was repeated for 45 cycles at 95°C for 5 seconds, and at 50°C for 30 seconds. One triplex PCR was used to quantify PMMoV, BRSV, and N1 (nucleocapsid gene) of SARS-CoV-2, while mtDNA was quantified in a singleplex assay. For the triplex PCR assay, each 40-µL reaction contained 10 µL of 4× One-step Advanced Probe Master Mix, 0.4 µL of 100× OneStep RT Mix, 2 µL of each 16× primer-probe mix of PMMoV, BRSV, and N1, 5 µL of nucleic acid extract, and 18.6 µL of RNase-free water. For the mtDNA singleplex master mix, each 40-µL reaction contained 10 µL of 4× One-step Advanced Probe Master Mix, 0.4 µL of 100× OneStep RT Mix, 2 µL of 16× primer-probe mix of mtDNA, 5 µL of 1:100 diluted nucleic acid extract, and 22.6 µL of RNase-free water. The master mix was pipetted into a QIAGEN QIAcuity 24 well 26k partition nanoplate.

### 2.4. Data Analysis and Statistics

We performed data analysis using R program (version 4.0.1). A logistic regression model was used to study the association between SARS-CoV-2 positivity and other variables including inpatient, outpatient, days after COVID-19 was confirmed, PMMoV, and mtDNA concentration. The Pearson correlation was used to estimate the association between log10-transformed PMMoV and mtDNA concentrations.

## 3. RESULTS

### 3.1. Participant Description

A total of 42 subjects with confirmed COVID-19 were enrolled in this study, and 155 fecal samples were collected throughout the 42-day follow-up period. Most of these subjects, 61.9% (26/42), were inpatients, and 38.1% (16/42) were outpatients. Most participants manifested severe clinical symptoms if hospitalized (inpatients), while outpatient participants usually had mild symptoms. The self-reported COVID-19 vaccination status indicated that 66.7% (28/42) were vaccinated (received at least one dose of COVID-19 vaccine and had a breakthrough infection) and 33.3% (14/42) were unvaccinated. Among these subjects, only 14.3% (6/42) were ≥65 years old (Table 1).

**Table 1.**
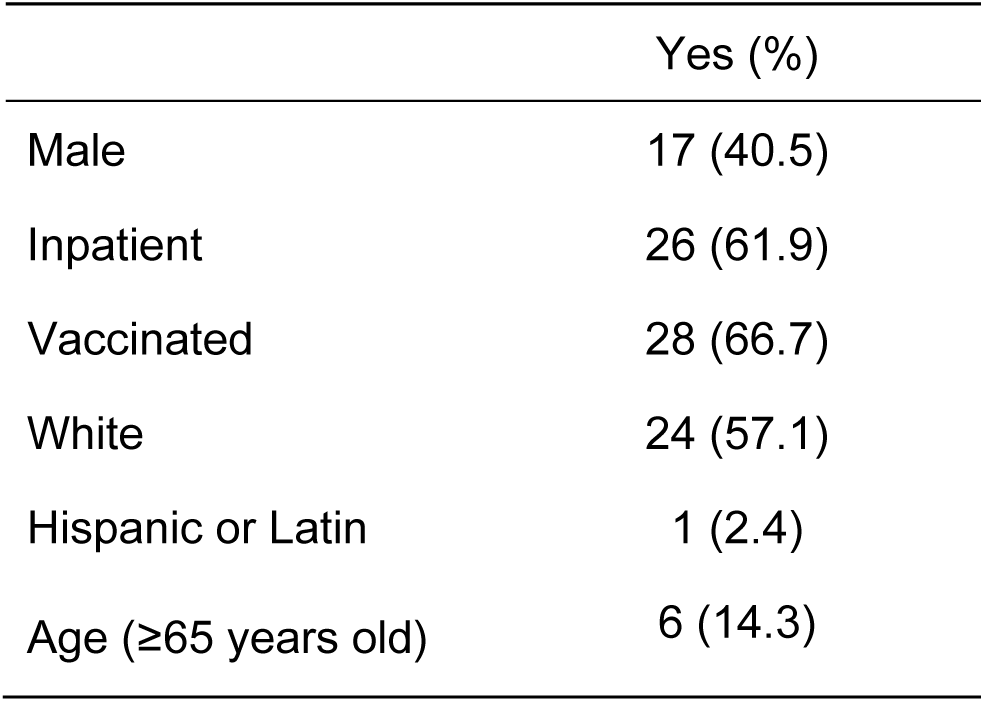
Demographic Characteristics of 42 Study Participants.

|  | Yes (%) |
| --- | --- |
| Male | 17 (40.5) |
| Inpatient | 26 (61.9) |
| Vaccinated | 28 (66.7) |
| White | 24 (57.1) |
| Hispanic or Latin | 1 (2.4) |
| Age (≥65 years old) | 6 (14.3) |

### 3.2. COVID-19 Clinical Symptoms

To compare the clinical course of infections of vaccinated and unvaccinated participants, clinical symptoms relevant to COVID-19 infection were assessed. Cough, diarrhea, fever, loss of smell, and shortness of breath were more frequently observed in unvaccinated subjects than in vaccinated subjects. Frequencies of headache and myalgia fatigue were higher in vaccinated and in unvaccinated groups. When inpatients and outpatients were compared, inpatients were much more likely to report cough, diarrhea, and fever than outpatients while outpatients tended to have more symptoms of headache, loss of smell, myalgia and fatigue. These differences may be caused by the small number of subjects in both groups (Table 2).

**Table 2.** Symptoms Among Patients Participants.

| Symptom | Vaccination Status |  | Admission Status |  |
| --- | --- | --- | --- | --- |
|  | Vaccinated (N=28) % | Unvaccinated (N=14) % | Inpatient (N=26) % | Outpatient (N=16) % |
| Cough | 35.7 | 71.4 | 61.9 | 18.8 |
| Diarrhea | 7.1 | 28.6 | 14.3 | 6.2 |
| Fever | 28.6 | 42.9 | 47.6 | 12.5 |
| Headache | 35.7 | 21.4 | 19.0 | 43.8 |
| Loss of smell | 17.9 | 21.4 | 19.0 | 31.2 |
| Loss of taste | 14.3 | 14.3 | 19.0 | 25.0 |
| Myalgia/fatigue | 60.7 | 50.0 | 47.6 | 75.0 |
| Shortness of Breath | 46.4 | 57.1 | 52.4 | 50.0 |
| Other symptoms | 21.4 | 0 | 23.8 | 18.8 |
| ICU* stay | 4.0 | 0 | 4.8 | 0 |

\*Intensive care unit

### 3.3. Longitudinal SARS-CoV-2 Fecal Shedding

The SARS-CoV-2 fecal shedding patterns for each inpatient and outpatient study subject are shown in Figure 1. SARS-CoV-2 RNA was detected in at least one of the fecal samples collected during the sampling period for 73.1% (19/26) of inpatient participants. Most SARS-CoV-2 RNA-positive samples were collected within two weeks of COVID-19 diagnosis. Five inpatients still shed the virus in their stools after two weeks, and one inpatient had detectable SARS-CoV-2 RNA in a stool sample at 59 days after COVID-19 diagnosis. For the 71 fecal samples collected from inpatients, 47.9% (34/71) had SARS-CoV-2 N1 gene measurements above the limit of detection of our PCR assay. In contrast, outpatients showed a different fecal shedding pattern. Only 7.1% of 84 fecal samples collected from 16 outpatients were SARS-CoV-2 RNA positive, and all of these came from four study subjects. The low proportion of SARS-CoV-2 positive stool samples from outpatients may be due to delays in sample collection. Interestingly, all four outpatient subjects with positive SARS-CoV-2 stool samples were intermittent shedders and were excreting the virus at three or more weeks after infection was confirmed.

**Figure 1.**
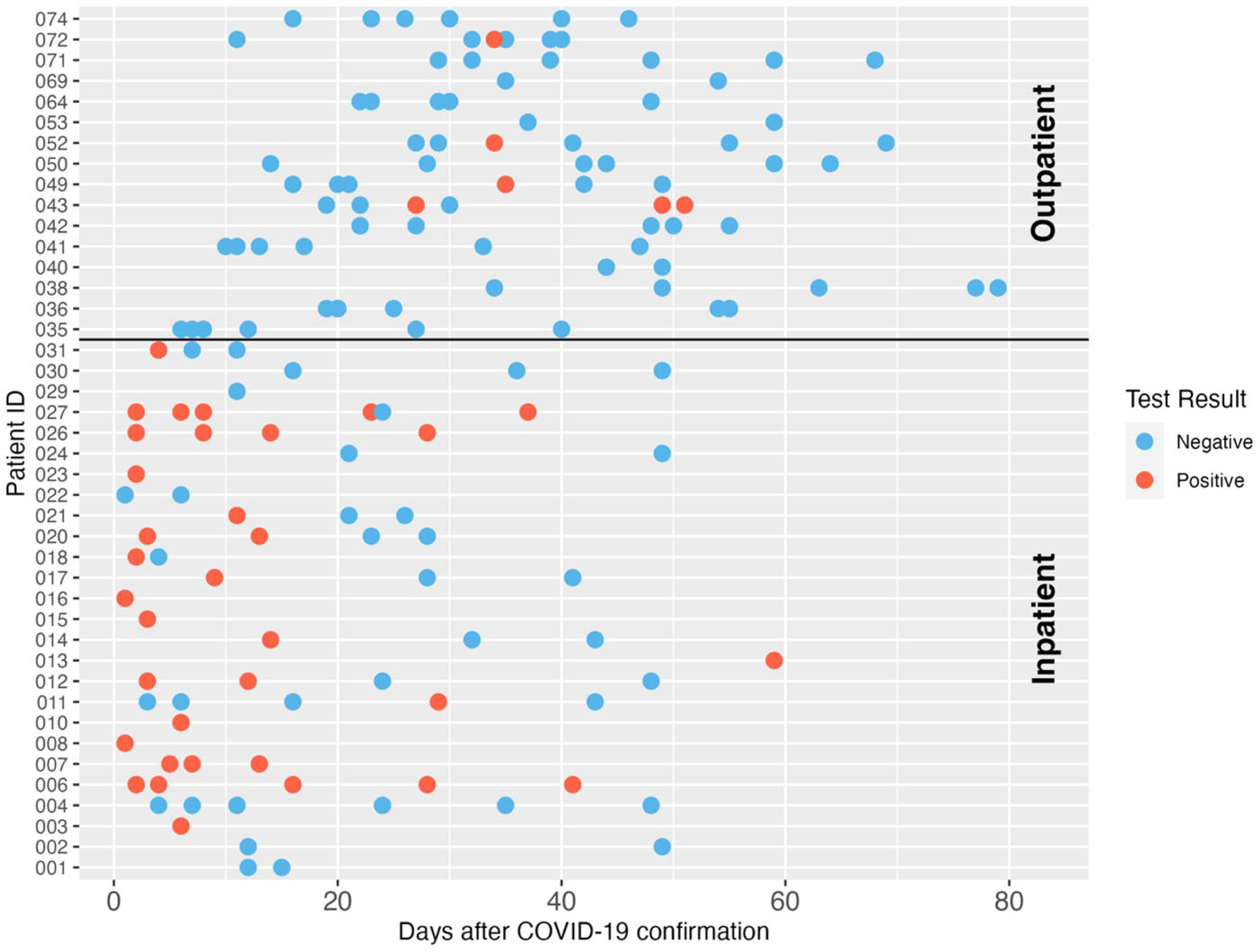
SARS-CoV-2 RNA longitudinal shedding dynamics in stool samples of COVID-19 patients in this study. The patient IDs are shown on the *y*-axis. The x-axis indicates days after COVID-19 diagnosis was confirmed. Patients 001–031 were enrolled as inpatients, and patients 035–074 were enrolled as outpatients. Three patients with unclear confirmation dates were excluded from this figure. Red circles indicate positive SARS-CoV-2 detection, and blue circles indicate the sample was negative for SARS-CoV-2 RNA by dPCR.

### 3.4. SARS-CoV-2 RNA Concentrations in Fecal Specimens

Concentrations of SARS-CoV-2 RNA in stool samples ranged from 4.5×10^3^ genome copies (GC)/dry gram to 1.19×10^9^ GC/dry gram with a median of 1.08×10^5^ GC/dry gram. For the 34 fecal samples from inpatients that tested positive for SARS-CoV-2 RNA, the median concentration of SARS-CoV-2 RNA was 1.35×10^5^ GC/dry gram. For the six fecal samples from outpatient participants that tested positive for SARS-CoV-2 RNA, the median concentration of SARS-CoV-2 RNA was 9.11×10^3^ GC/dry gram (Figure 2, bottom left). Both the SARS-CoV-2 detection rates in stool samples and the mean virus RNA log10 concentrations in stool were significantly different in samples from inpatients versus outpatients (p<0.001 and p=0.003, respectively).

**Figure 2.**
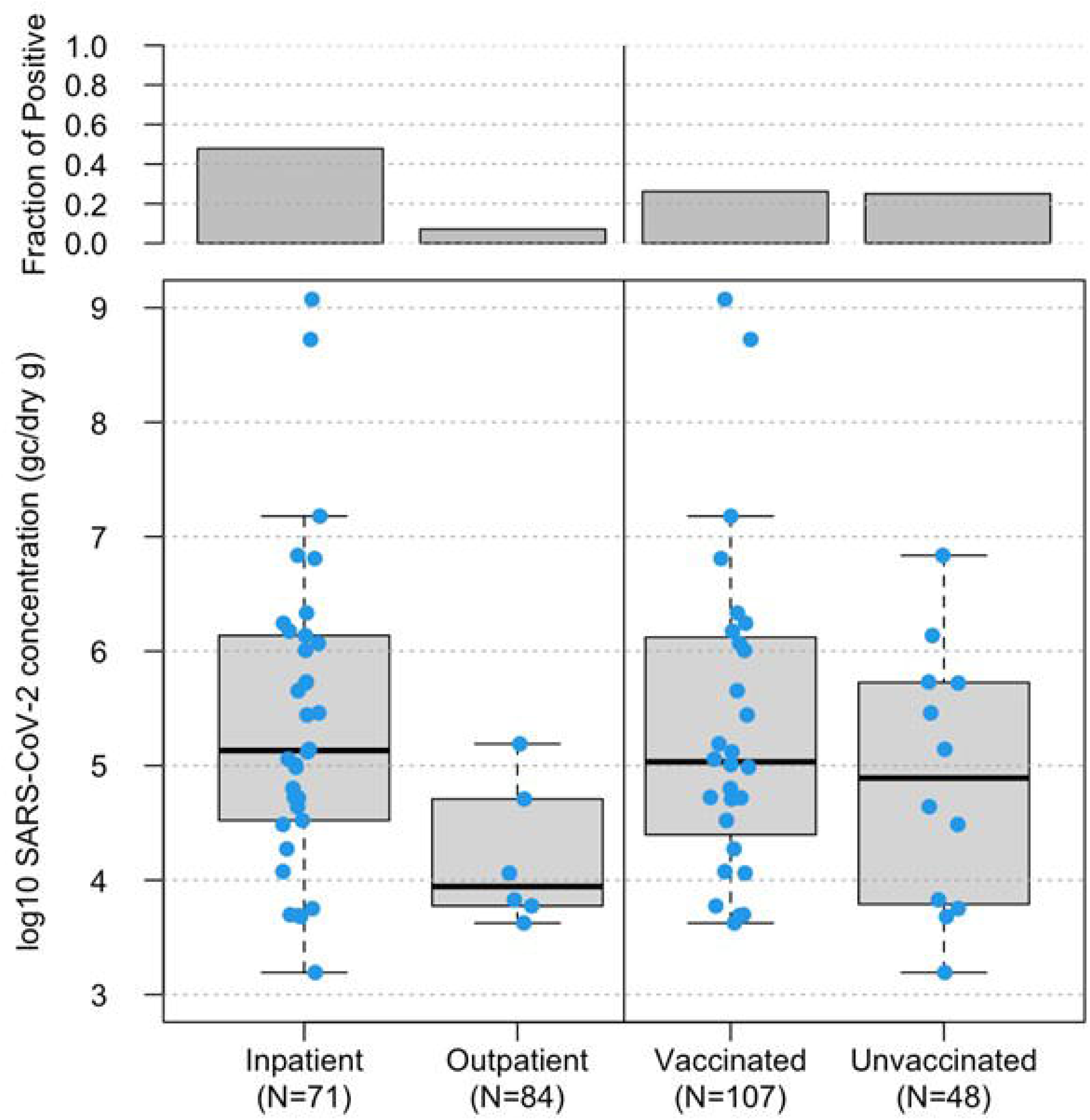
Boxplot of log_10_-transformed SARS-CoV-2 RNA concentrations (genome copies/dry gram, *y*-axis) in 155 fecal samples from inpatients vs. outpatients, based on status when enrolled, (bottom left, *x*-axis) and 146 fecal samples from vaccinated vs. unvaccinated study subjects (bottom right, *x*-axis). The horizontal lines in boxes denote the 50^th^ percentiles of SARS-CoV-2 RNA concentrations. The top boxes represent the fraction of SARS-CoV-2 RNA positive samples in each group.

### 3.5. Fecal Shedding Among Vaccinated (Breakthrough) and Unvaccinated Participants

There was little difference in the detection rates and concentrations of SARS-CoV-2 RNA in fecal samples from study subjects with different COVID-19 vaccination status. For the 28 vaccinated participants, 53.6% (15/28) of vaccinated participants shed SARS-CoV-2 in at least one of the fecal specimens collected during the study period, and 26.2% (28/107) fecal samples from vaccinated participants were SARS-CoV-2 RNA positive with a median concentration of 1.08×10^5^ GC/dry gram. There were 14 participants who reported that they were unvaccinated, and 8 of them (57.1%) had evidence of SARS-CoV-2 shedding in at least one of the collected fecal samples. Of the 48 fecal samples collected from unvaccinated participants, 25.0% (12/48) fecal samples from unvaccinated participants were positive for SARS-CoV-2 RNA and the median concentration was 9.14 ×10^4^ GC/dry gram (Figure 2, bottom right).

### 3.6. PMMoV Detection in Fecal Samples

PMMoV was analyzed in each fecal sample by dPCR on the same day as the SARS-CoV-2 dPCR to observe the biological variability of this fecal indicator within and between individuals over the sampling period. Almost all (99.4%, 154/155) of the fecal samples in this study had detectable PMMoV. The median concentration of PMMoV RNA was 1.73×10^7^ GC/dry gram. PMMoV RNA concentrations were highly variable over the sampling period within and between the individual study subjects. Some individuals showed relatively consistent concentrations of PMMoV RNA in their longitudinal samples; while for others the PMMoV RNA concentrations varied across several orders of magnitude among all the samples from an individual. The lowest PMMoV concentration was 790 GC/dry gram, and the highest concentration was 3.51×10^9^ GC/dry gram (Figure 3).

**Figure 3.**
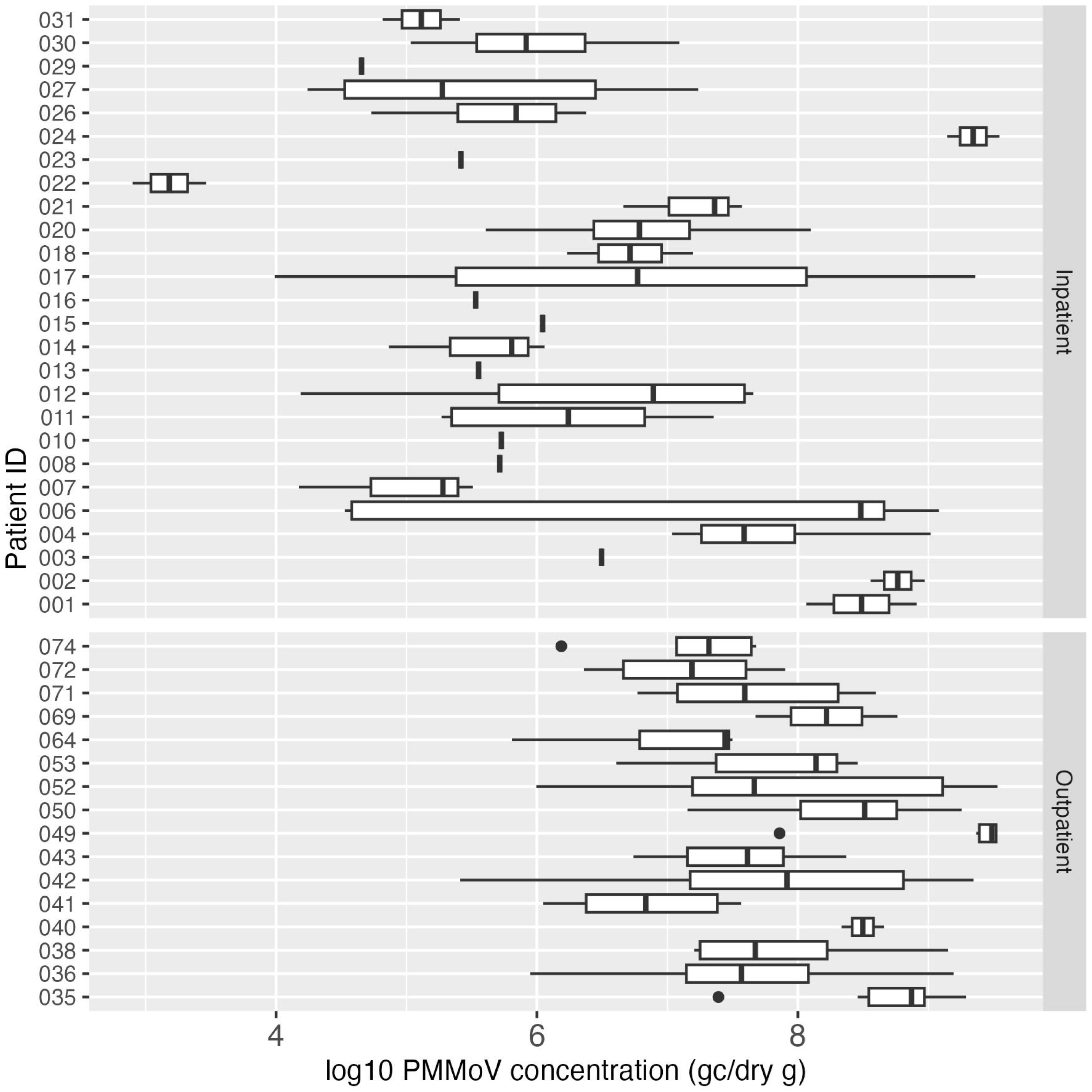
Boxplot of log_10_-transformed PMMoV concentration (genome copies/dry gram) in inpatient (top) and outpatient (bottom) study subjects. Each box summarizes the range of PMMoV concentrations in multiple fecal samples from an individual, and the vertical line in the box represents the median PMMoV concentration. The vertical line without the box indicates only one sample was tested for PMMoV for that individual.

### 3.7. Human mtDNA Detection in Fecal Samples

Human mtDNA, another fecal indicator, was analyzed in all the collected fecal samples. All 155 samples (100%) tested positive for mtDNA, and the median concentration of mtDNA in all positive samples was 2.49×10^8^ GC/dry gram. The concentration of mtDNA ranged between 8.22×10^6^ GC/dry gram and 3.88×10^10^ GC/dry gram. In contrast to PMMoV, the mtDNA concentration in stool was more consistent both between and within the study subjects over the study period (Figure 4).

**Figure 4.**
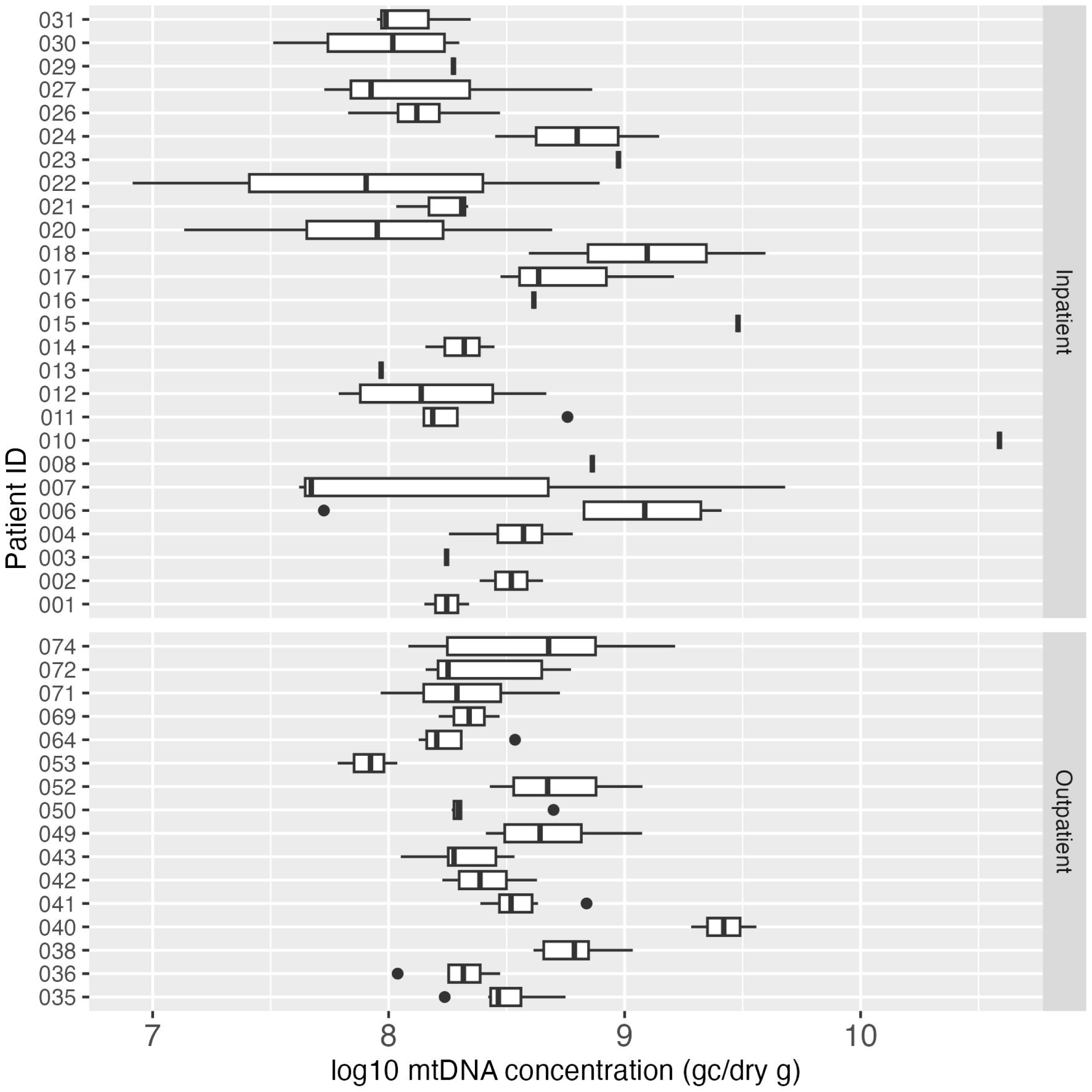
Boxplot of log_10_-transformed mtDNA concentration (GC/dry gram) in inpatient (top) and outpatient (bottom) study subjects. Each box summarizes the range of mtDNA concentrations in multiple fecal samples from an individual, and the vertical line in the box represents the median mtDNA concentration. The vertical line without the box indicates only one sample was tested for mtDNA for that individual.

### 3.8. Correlation between PMMoV and mtDNA

Pearson correlation analysis was performed to examine the association between PMMoV and mtDNA concentrations, the two human fecal markers measured in the fecal samples in this study. Figure 5 indicated a weak correlation (p = 0.16) between the concentrations of these two markers. Interestingly, mean log10 PMMoV concentration was significantly higher (p < 0.001) in fecal specimens from outpatients compared to those from inpatients.

**Figure 5.**
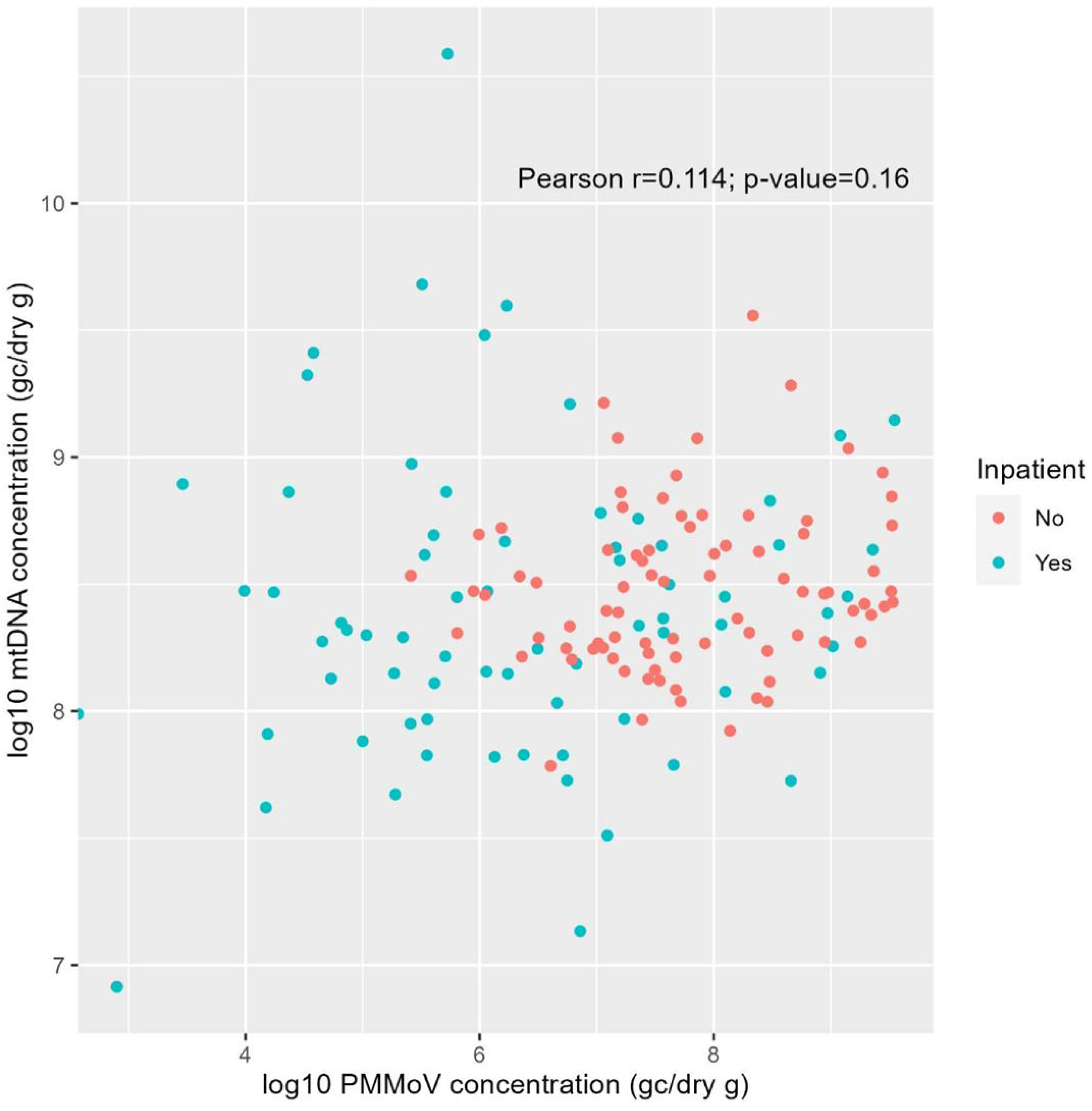
Pearson correlation between log_10_-transformed PMMoV RNA and mtDNA concentrations (GC/dry gram) from fecal samples of inpatients (green) and outpatients (red) in this study.

## 4. DISCUSSION

In this study, we describe frequency, duration, and concentration of SARS-CoV-2, PMMoV, and mtDNA shedding in fecal specimens from inpatient and outpatient study subjects with COVID-19 over a 42 day period after the confirmation of SARS-CoV-2 infection. Consistent with other reports (24–26), study subjects who had been vaccinated but had breakthrough infections were less likely to exhibit some clinical symptoms, such as cough, diarrhea, fever, loss of smell, loss of taste, and shortness of breath, compared to unvaccinated study subjects. Inpatient participants were more likely to be shedding SARS-CoV-2 RNA in their fecal specimens and had higher concentrations of the virus in their stool compared to outpatient participants. These results provide important information about the dynamics of the fecal shedding of SARS-CoV-2 and two human fecal indicators. Understanding the presence, magnitude, and duration of these targets of interest is critical for the broad application of WBE and interpretation of WBE results.

Although several cross-sectional studies have reported SARS-CoV-2 fecal shedding in patients with COVID-19 at the early stage of infection, little is known about longitudinal SARS-CoV-2 shedding. In this study, we observed that 73.1% (19/26) inpatients shed SARS-CoV-2 up to three weeks and 19.2% (5/26) inpatients continued to shed viruses beyond day 20; however only 25.0% (4/16) outpatients shed viruses after three weeks of diagnosis. In contrast, the study led by Natarajan et al.,(3) included a total of 113 individuals diagnosed with COVID-19 who were followed for up to 10 months. They reported that 49.2% of fecal specimens tested positive for SARS-CoV-2 during the week following diagnosis using PCR-based methods, with the positivity rate declining to 12.7% at 4 months and 3.8% at 7 months after diagnosis. Another study reported the fecal shedding results from 48 SARS-CoV-2 infected individuals and observed that approximately 80% of the fecal samples collected within the first 5 days were positive for SARS-CoV-2, and this positivity rate dropped to 10% at 28 days after symptom onset. A median concentration of three orders GC of magnitude/mg dry weight was reported in positive fecal samples (27), which is one order GC of magnitude higher compared to this study.

Normalizing SARS-CoV-2 concentrations measured in wastewater with a human fecal indicator is a common practice in order to adjust for factors that may contribute to the variability in SARS-CoV-2 concentrations from distinct catchment areas and the recovery of the virus from wastewater with different methods. Several fecal indicators, including human ribonuclease P (37), PMMoV (38), Bacteroides HF183 (39); F-specific RNA bacteriophages (40), human 18S rRNA (41), cross-assembly phage (CrAssphage) (42), have been proposed. Among these recommended fecal indicators, PMMoV is widely used. PMMoV is a plant RNA virus associated with pepper products and human diet (17, 38). The presence and magnitude of PMMoV RNA in biological and environmental samples are varied due to the geographic and dietary variations between populations and individuals. These variations may complicate the application of PMMoV as a normalizing indicator in some situations, such as countries with less consumption of plants and pepper products. Although PMMoV has been consistently detected in wastewater and has been proposed as a normalization indicator, there are few reports characterizing PMMoV detection in human fecal samples. Additionally, high concentrations of PMMoV have been detected in non-human fecal samples, such as chickens and seagulls (43, 44), indicating that PMMoV is not human-specific and detection of PMMoV in wastewater may originate from other non-human sources, eg., livestock and wildlife. Therefore, accurate characterization of the frequency of detection and quantification of PMMoV in human fecal samples provides critical information for better application of this marker as a normalization control in WBE. In this study, PMMoV was detected in 99.4% fecal samples, with a median concentration of 1.73×10^7^ GC/dry gram. The PMMoV concentrations measured in this study are similar to those reported in one recent publication that also quantified PMMoV in human stool samples using digital PCR and reported dry mass concentrations (27). In concordance with their results, we observed significant variations of PMMoV concentrations between and within individuals throughout the sampling period. This suggests that PMMoV levels in human fecal materials may be affected by an individual’s daily diet and lifestyle.

Human mtDNA is a human-specific intrinsic genetic marker for fecal source tracking. This marker is abundant in human feces and sewage which makes it a useful indicator of human fecal contamination for environmental microbial research and risk assessment applications (22, 23, 45, 46). There are limited reports on the use of mtDNA as a normalization control in SARS-CoV-2 wastewater studies. In this study, mtDNA was detected in 100% fecal samples, with a median concentration of 2.49×10^8^ GC/dry gram. Our mtDNA concentrations are within the range of mtDNA concentrations in human feces reported by other studies(45). Compared to PMMoV detection in human feces, mtDNA was detected in all the fecal specimens, was present at ten-fold higher concentrations, and exhibited little variability between and within individuals. Furthermore, there are no non-human sources of this marker. These characteristics suggest that mtDNA may be better suited for use as a normalization factor for WBE results than PMMoV.

There are several limitations in this study. First, the sample size was small, with 42 confirmed COVID-19 patients enrolled and 155 fecal samples collected. This population was not large enough to allow for stratification for further examination of the effects of some demographic variables on the fecal shedding of SARS-CoV-2. Second, the recruited subjects were from a limited geographic area, basically within the metro Atlanta area of Georgia. Therefore, the sample may not be representative of the US population, and the conclusions may not be generalizable. Third, the fecal samples were collected between March 21, 2021 and July 28, 2022, when the Delta and Omicron variants were prevalent. We do not know the shedding dynamics of other SARS-CoV-2 variants, and the small sample size did not allow us to compare the shedding dynamics of the Delta versus Omicron variants. Fourth, outpatient participants had much lower proportion of SARS-CoV-2 positive fecal specimens than inpatient participants in this study, and surprisingly, all the fecal samples from outpatient study subjects were negative for SARS-CoV-2 within the first three weeks of COVID-19 diagnosis. The low SARS-CoV-2 detection rate in fecal specimens from outpatient participants may be due to the delays between subject enrollment and stool sample collection and possible mishandling during storage and shipment from the participants’ homes to the research laboratory. In contrast, fecal specimens from inpatient participants were collected earlier in the course of infection and stored under optimum conditions until analysis. Despite these limitations, the quantitative measurements of SARS-CoV-2, PMMoV, and mtDNA in longitudinal fecal samples from confirmed COVID-19 patients have significant relevance to our understanding of COVID-19 epidemiology, and the SARS-CoV-2 shedding information addresses a critical knowledge gap for the advancement of WBE and the use of wastewater monitoring data for SARS-CoV-2 to estimate COVID-19 prevalence and incidence in specific catchment populations.

## Data Availability

All data produced in the present study are available upon reasonable request to the authors

## ACKNOWLEDGEMENTS

This study was supported by the NIH Rapid Acceleration of Diagnostics (RADx) initiative (contract No. 75N92021C00012 to Ceres Nanosciences, Inc) through a contract from Ceres Nanosciences to Emory University. We are grateful to Salvins J. Strods, Shawn Mulvaney, Chipo Chemoyo J. Baker Afamefuna, and Jenica Patterson with the NIH RADx Initiative, and Ross Dunlap, Robbie Barbero, and Ben Lepene from Ceres Nanosciences, Inc.

